# Plasma biomarkers identify brain ATN abnormalities in a dementia-free population-based cohort

**DOI:** 10.1101/2025.04.27.25326360

**Authors:** Menayit Tamrat Dresse, Pamela C. L. Ferreira, Akshay Prasadan, Jihui L. Diaz, Xuemei Zeng, Bruna Bellaver, Guilherme Povala, Victor L. Villemagne, M. Ilyas Kamboh, Ann D. Cohen, Tharick A. Pascoal, Mary Ganguli, Beth E. Snitz, C. Elizabeth Shaaban, Thomas K. Karikari

**Affiliations:** Department of Epidemiology, School of Public Health, University of Pittsburgh, Pittsburgh, PA 15213, USA; Department of Psychiatry, School of Medicine, University of Pittsburgh, 3811 O’Hara Street, Pittsburgh, PA 15213, USA; Department of Statistics & Data Science, Dietrich College of Humanities and Social Sciences, Carnegie Mellon University, Pittsburgh, PA, 15213, USA; Alzheimer’s Disease Research Center, University of Pittsburgh, Pittsburgh, PA, USA; Department of Human Genetics, School of Public Health, University of Pittsburgh, Pittsburgh, PA 15213, USA; Department of Neurology, School of Medicine, University of Pittsburgh, Pittsburgh, PA 15213, USA

**Author notes:** Corresponding author: Department of Psychiatry, School of Medicine, University of Pittsburgh, Pittsburgh 15213, PA, USA. Joint senior authors (C. Elizabeth Shaaban, Thomas K. Karikari).

**Keywords:** Amyloid beta, tau pathology, neurodegeneration, neuroimaging, ATNframework, p-tau217, p- tau181, p-tau231, GFAP, neurofilament light chain

## Abstract

**INTRODUCTION:** Using the ATN framework, we evaluated the potential of plasma biomarkers to identify abnormal brain amyloid-beta (Aβ) positron emission tomography (PET), tau-PET and neurodegeneration in a socioeconomically disadvantaged population-based cohort.

**METHODS:** Community-dwelling dementia-free (n=113, including 102 (91%) cognitively normal) participants underwent ATN neuroimaging and plasma biomarker assessments.

**RESULTS:** Plasma Aβ42/Aβ40, p-tau181, and p-tau217 showed significant associations with Aβ-PET status (adjusted odds ratio [AOR] of 1.74*10^-24^, 1.47, and 3.43*10^3^ respectively [p-values<0.05]), with p-tau217 demonstrating the highest classification accuracy for Aβ-PET status (AUC=0.94). Plasma p-tau181 and p-tau217 showed significant associations with tau-PET status (AOR: 1.50 and 22.24, respectively, p-values<0.05), with comparable classification accuracies for tau-PET status (AUC=0.74 and 0.70, respectively). Only plasma NfL showed significant association with neurodegeneration based on cortical thickness (AOR=1.09, p-value<0.05).

**CONCLUSION:** Our findings highlight potential of plasma p-tau217 as a biomarker for brain Aβ and tau pathophysiology, p-tau181 for tau abnormalities, and NfL for neurodegeneration in the community.

## 1. Background

Currently, there is no known cure for Alzheimer’s disease (AD) [1]. However, disease-modifying treatments approved by the US Food and Drug Administration (FDA) that can help remove brain amyloid-beta (Aβ) aggregates and slow disease progression have recently become available [1–3]. Given this, accurate biomarker-supported assessment during the early stages of the AD continuum is essential for timely and effective intervention [4].

AD has long been clinically diagnosed at a relatively late stage of the biological disease, once overt symptoms of dementia have developed, hindering early detection [5]. The 2018 National Institute of Aging-Alzheimer’s Association (NIA-AA) AD research framework, updated and published in June 2024 [6], greatly shifted this syndromic approach to a biological (etiological) diagnosis [7]. This framework groups the main pathological hallmarks of AD under amyloidosis (A), tau pathology (T) and neurodegeneration (N), using validated cerebrospinal fluid (CSF), positron emission tomography (PET) and structural magnetic resonance imaging (MRI) biomarkers [7]. Despite their proven diagnostic importance, the somewhat invasive nature of CSF collection and the high cost and limited accessibility of PET imaging restrict their use on a large scale in routine clinical settings and applicability for repeated population-wide evaluation.

In recent years, more accessible and cost-effective diagnostic approaches, such as plasma biomarkers, have been widely developed and evaluated as promising surrogate markers and predictors of Aβ and tau pathologies, and neurodegeneration [6]. Many of these are included in the 2024 revised criteria of AD diagnosis and staging by the Alzheimer’s Association (AA) workgroup [6]. Plasma Aβ42/Aβ40, one of the core Aβ proteinopathy biomarkers, shows a strong negative correlation with brain Aβ pathology measured by PET, especially in the pre-clinical stage of AD [8,9]. Plasma glial fibrillary acidic protein (GFAP), an intermediate filament protein and a marker of astrocytosis [10], significantly associates with brain Aβ signals and predict Aβ-PET abnormalities [11,12]. In the 2024 revised criteria, GFAP is categorized under “inflammation” representing a biomarker of a non-specific process in AD pathophysiology [6]. Several plasma phosphorylated-tau (p-tau) variants have been developed and validated as candidate biomarkers of both Aβ and tau pathologies of which p-tau181, p-tau217 and p-tau231 are the most studied [12–27]. For neurodegeneration, plasma neurofilament light chain (NfL), a neuronal cytoplasmic protein, has been identified as a reliable, albeit non-specific, marker of brain atrophy [28,29]. Plasma NfL associates well with neuroimaging measures of neurodegeneration, although it lacks specificity for underlying AD-related pathology [28].

However, several of these studies reported individual plasma biomarkers as predictors of Aβ, tau or neurodegeneration abnormalities, often lacking head-to-head evaluation. Additionally, participants involved in such studies were usually recruited from urban academic medical settings and tended to have high socioeconomic status (SES) [30]. This recruitment approach overlooks populations living in medically underserved areas and with low SES, who face significant dementia-related disparities [31–33] resulting in limited external validity. The emerging interdisciplinary population neuroscience approach addresses this challenge by emphasizing the need for studying biofluid and neuroimaging biomarkers within population-based cohorts [34]. This approach enables the identification of biomarker trajectories at the population level while accounting for factors, including SES, that may influence expression and predictive values of the biomarkers [34]. Thus, such population-based studies of AD plasma biomarkers ensure external validity (generalizability) of findings, thereby enriching our understanding of AD in the broader community.

Therefore, in this study our aims were: (i) to evaluate associations in parallel between six plasma biomarkers (Aβ42/Aβ40, GFAP, NfL, p-tau181, p-tau217 and p-tau231) and brain Aβ- PET, tau-PET and magnetic resonance imaging (MRI)-based neurodegeneration measures; and (ii) to assess the accuracy of each plasma biomarker in classifying brain Aβ-PET, tau-PET and neurodegeneration status, in the Monongahela Youghiogheny Healthy Aging Team-Neuroimaging (MYHAT-NI) study. The MYHAT-NI study collected neuroimaging data from a subset of participants enrolled in the large population-based prospective cohort study, MYHAT [35]. Participants in the parent study were recruited from a small-town area in southwestern Pennsylvania, with some areas medically underserved. This region, once a hub of steel manufacturing, experienced regional economic decline following the collapse of the industry in the late 1970s and 1980s [35,36].

## 2. Methods

2.1. **Study participants**

We included 113 participants from the MYHAT-NI study, a neuroimaging study which enrolled a subset of participants from the parent MYHAT cohort. The MYHAT study is an ongoing population-based study of dementia-free older adults drawn from a Rust Belt region of southwestern Pennsylvania, United States of America (USA) [36]. Participants aged 65 years or older and not living in a long-term care institution were randomly selected for recruitment from voter registration lists of selected towns in a geographically defined region. Participants were excluded if they were too ill to participate, had severe cognitive impairment, significant vision or hearing impairment, or were decisionally incapacitated. Inclusion criteria for the MYHAT-NI study were participation in the parent MYHAT study and having a Clinical Dementia Rating (CDR) sum-of-boxes score, the sum of ratings in each six cognitive domains assessed in the CDR evaluation [37], <1.0. The only exclusion criteria were contraindication for MRI or PET neuroimaging. All participants gave written informed consent, and all study procedures were approved by the Institutional Review Board of the University of Pittsburgh. Further details on recruitment and assessment procedures have been described in detail previously [35,36].

### 2.2. Neuroimaging methods

Neuroimaging was conducted at the time of blood collection at the University of Pittsburgh PET Center. T1-weighted magnetization prepared rapid gradient echo (MPRAGE) structural MRI series were obtained for each participant using a 3T Siemens PRISMA scanner before the PET imaging session. All PET images were acquired using a Siemens Biograph mCT Flow 64-4R PET/CT scanner.

[^11^C]Pittsburgh Compound B (PiB) (15 mCi) for Aβ pathology imaging or [^18^F]AV-1451 (7–10 mCi) for tau pathology imaging were administered as slow bolus injections via the antecubital vein. PET emission data were collected in a series of 5-minute frames spanning 50–70 min post-injection for [^11^C]PiB and 80–100 min for [^18^F]AV-1451. [^11^C]PiB and [^18^F]AV-1451 acquisition frames were summed into a single 20-minute frame and registered to the participant’s T1 MR image using the normalized mutual information algorithm [35].

Participants’ neuroimaging-based “A”, “T”, and “N” statuses were classified based on [^11^C]PiB, [^18^F]AV-1451 and MRI scans. Neocortical Aβ load was calculated as the mean ^11^C]PiB Standard Uptake Value Ratio (SUVR) of nine composite regions: anterior cingulate, posterior cingulate, ventral striatum, superior frontal cortex, orbitofrontal cortex, insula, lateral temporal cortex, parietal, and precuneus [38]. For tau load, a composite [^18^F]AV-1451 SUVR was computed by normalizing the meta temporal composite region (comprising amygdala, entorhinal, fusiform, parahippocampal, inferior temporal and middle temporal) to FreeSurfer cerebellar gray matter as a reference [39]. MRI-based cortical thickness composite measure was derived from a surface-area weighted average of the mean cortical thickness of four FreeSurfer regions of interest (ROIs) that are most predictive of AD pathology: entorhinal, inferior temporal, middle temporal, and fusiform [39,40].

Participants were classified as Aβ-PET positive (A+) or Aβ-PET negative (A-) based on a pre- defined [^11^C]PiB global SUVR cutoff of >1.346 as A+, and [^11^C]PiB global SUVR <=1.346 as A- [41–43]. Participants with [^18^F]AV-1451 meta temporal SUVR >1.18 were considered tau-PET positive (T+), while those with a [^18^F]AV-1451 meta temporal SUVR <= 1.18 were considered tau-PET negative (T−) [39]. Participants with cortical thickness composite measure <2.7 were classified as neurodegeneration positive (N+) while those with cortical thickness composite measure >= 2.7 were classified as neurodegeneration negative (N-) [40]. Full neuroimaging details were described previously [35].

### 2.3. Plasma biomarker measurements

All plasma biomarkers in this study were measured using Single molecule array (SIMOA) methods on the HD-X instrument (Quanterix, Billerica, MA, USA) at the Department of Psychiatry, School of Medicine, University of Pittsburgh, USA, [44], except for p-tau231 which was measured at the Clinical Neurochemistry Laboratory, Sahlgrenska University Hospital, Mölndal, Sweden. All frozen samples were thawed at room temperature and centrifuged at 4000xg for 10 min to remove particulates prior to the measurements. Plasma Aβ42, Aβ40, GFAP and NfL were measured with the Neurology 4-Plex E (#103670). P-tau181 was measured with V2 Advantage kit (#103714); p-tau217 with the ALZpath Simoa® p-tau217 V2 Assay Kit (#104371); and p-tau231 using *in-house* SIMOA methods, described previously [21].

For each assay, two or three quality control (QC) samples of different concentrations were analyzed in duplicate both at the start and end of each technical run to assess reproducibility of each assay. The average within-run coefficients of variation (CVs) of the QC samples were 8.9% for Aβ42, 9.5% for Aβ40, 9.9% for GFAP, 14.3% for NfL 6.6% for p-tau181 and 3.7% for p- tau217. The average between-run CVs were 13.0% for Aβ42, 14.6% for Aβ40, 17.8% for GFAP, 18.3% for NfL, 11.7% for p-tau181, and 11.4% for p-tau217 [44].

### 2.4. Apolipoprotein E genotyping

Apolipoprotein E (*APOE*) genotyping was carried out using blood or saliva samples as described previously [45]. For this analysis, individuals with one or two *APOE4* allele were grouped into *APOE4* carriers and individuals without an *E4* allele were grouped as non- *APOE4* carriers.

### 2.5. Statistical analysis

Continuous variables were summarized using mean and standard deviation (SD) and categorical variables and complete neuroimaging-based ATN profile distribution of the study participants were reported using count and percentage. The difference in demographics and plasma biomarkers between neuroimaging-based ATN statuses was tested using Wilcoxon rank-sum test and Fisher’s exact test for continuous and categorical variables, respectively. Plasma biomarker concentrations (pg/mL) were not normally distributed, and thus they were log transformed. Box plots of the distributions of the log-transformed plasma biomarkers by neuroimaging-based ATN statuses were inspected.

The classification accuracies of the AD plasma biomarkers were tested, with neuroimaging-based ATN biomarkers serving as the ground truth. A series of logistic regression models using complete data were fitted to examine this, and both unadjusted and adjusted odds ratios (OR) and their 95% confidence intervals (CI) are reported. Adjusted logistic regression models were controlled for age, sex, education level, *APOE4* carrier status and CDR score. A two-sided p-value less than 0.05 was considered statistically significant. Multiple comparisons correction was not applied in the logistic regression models. Receiver operating characteristics (ROC) curves and their respective areas under the curve (AUC) were estimated using values predicted by the logistic regression models to investigate the performance of these plasma biomarkers to predict ATN status.

The associations of Aβ-PET and tau-PET with plasma biomarkers were also tested using voxel-wise linear regressions. Models were adjusted for age, sex, education level, *APOE4* carrier status and CDR score. In the voxel-wise analyses, multiple comparisons correction was performed using random field theory (RFT), with a voxel threshold of P < 0.001. All analyses were conducted in R 4.1.1 (R foundation for Statistical Computing, Vienna, Austria; http://www.r-project.org/), and imaging analyses were carried out using the RMINC package.

Sensitivity analysis was also conducted to evaluate the impact of Body Mass Index (BMI) and Area Deprivation Index (ADI) national rank, a measure of neighborhood deprivation (higher is worse deprivation), on the associations of the plasma biomarkers with neuroimaging-based ATN biomarkers and their classification accuracy of neuroimaging-based ATN status. To evaluate this a series of logistic regression models were fitted by additionally controlled for BMI and ADI in the adjusted models.

## 3. Results

### 3.1. Characteristics of study participants

As shown in **Table 1**, n=113 dementia-free participants were included in the study. Based on neuroimaging biomarkers of ATN, 28 (25%) individuals were A+, 39 (36%) were T+ and 33 (29%) were N+. Distribution of the neuroimaging-based ATN profiles among the study participants is shown in **Supplementary Table 1**. The most prevalent profile was A-T-N- (40%) followed by A-T-N+ (17.7%), A+T+N- (13.3%) and A-T+N- (12.4%). The mean (standard deviation (SD)) age of the participants was 77 (6) years, a slight majority were female (54%), most identified as non-Hispanic White (95%), had education attainment of high school or less (57%), had high ADI (57%) (while the ADI of the study cohort range from 32–99) and 16% were *APOE*4 carriers. This study cohort comprised mostly cognitively normal individuals (91% with CDR=0) with a mean (SD) BMI of 28.0 (4.4).

**Table 1.** Study participants’ characteristics and biomarker profiles by neuroimaging-based ATN status groups.

|  |  | Aβ-PET status |  |  | Tau-PET status |  |  | Neurodegeneration status |  |  |
| --- | --- | --- | --- | --- | --- | --- | --- | --- | --- | --- |
|  | Total<br>N= 113 | A+<br>N= 28<br>(25%) | A-<br>N= 85<br>(75%) | p-<br>value<sup>a</sup> | T+<br>N= 39<br>(35%) | T-<br>N= 74<br>(65%) | p-<br>value<sup>a</sup> | N+<br>N= 33<br>(29%) | N-<br>N= 80<br>(71%) | p-<br>value<sup>a</sup> |
| **Demographic factors** |  |  |  |  |  |  |  |  |  |  |
| Age, Mean (SD) | 77 (6) | 79 (7) | 76 (6) | 0.055 | 77 (6) | 77 (6) | 0.9 | 80 (7) | 75 (5) | **0.002** |
| Sex, female, n (%) | 61 (54%) | 18 (64%) | 43 (51%) | 0.2 | 23 (59%) | 38 (51%) | 0.4 | 17 (52%) | 44 (55%) | 0.7 |
| Race/ethnicity, non-Hispanic White, n (%) | 107 (95%) | 27 (96%) | 80 (94%) | >0.9 | 38 (97%) | 69 (93%) | 0.7 | 32 (97%) | 75 (94%) | 0.7 |
| **Socioeconomic factor** |  |  |  |  |  |  |  |  |  |  |
| Education, n (%) |  |  |  | 0.14 |  |  | 0.6 |  |  | 0.6 |
| High School or Less | 64 (57%) | 15 (54%) | 49 (58%) |  | 23 (59%) | 11 (15%) |  | 20 (61%) | 44 (55%) |  |
| Partial/Full College/Trade | 35 (31%) | 12 (43%) | 23 (27%) |  | 13 (33%) | 22 (30%) |  | 8 (24%) | 27 (34%) |  |
| Graduate Degree | 14 (12%) | 1 (3.6%) | 13 (15%) |  | 3 (7.7%) | 11 (15%) |  | 5 (15%) | 9 (11%) |  |
| High Area Deprivation Index<sup>b</sup>, n(%) | 63 (57%) | 14 (52%) | 49 (59%) | 0.5 | 20 (53%) | 43 (60%) | 0.5 | 18 (56%) | 45 (58%) | 0.9 |
| **Genetic factor** |  |  |  |  |  |  |  |  |  |  |
| APOE4 carriers, n (%) | 18 (16%) | 12 (43%) | 6 (7.1%) | **<0.001** | 9 (23%) | 9 (12%) | 0.13 | 5 (15%) | 13 (16%) | 0.9 |
| **Cognitive function assessment** |  |  |  |  |  |  |  |  |  |  |
| CDR=0, n (%) | 102 (91%) |  |  |  |  |  |  |  |  |  |
| CDR=0 | 10 (9%) | 22 (79%) | 80 (95%) | **0.015** | 35 (90%) | 67 (92%) | 0.7 | 28 (85%) | 74 (94%) | 0.2 |
| CDR=0.5 |  | 6 (21%) | 4 (5%) |  | 4 (10%) | 6 (8%) |  | 5 (15%) | 5 (6%) |  |
| MMSE Score, n (%) | 108 (96%) |  |  |  |  |  |  |  |  |  |
| >= 24 | 4 (4%) | 27 (96%) | 81 (96%) | 0.4 | 38 (97%) | 70 (96%) | 0.4 | 32 (97%) | 76 (96%) | **0.025** |
| 19–23 |  | 1 (4%) | 3 (4%) |  | 1 (3%) | 3 (4%) |  | 1 (3%) | 3 (4%) |  |
| **Health-related factor** |  |  |  |  |  |  |  |  |  |  |
| Body Mass Index, Mean (SD) | 28.0 (4.4) | 27.0 (4.6) | 28.4 (4.2) | 0.2 | 27.6 (4.7) | 28.3 (4.2) | 0.6 | 28.5 (4.1) | 27.9 (4.5) | 0.5 |
| **Neuroimaging measures, Mean (SD)** |  |  |  |  |  |  |  |  |  |  |
| [<sup>11</sup>C]PiB global SUVR | 1.28 (0.31) | 1.74 (0.28) | 1.12 (0.07) | **<0.001** | 1.46 (0.40) | 1.18 (0.19) | **<0.001** | 1.26 (0.31) | 1.29 (0.31) | 0.4 |
| [<sup>18</sup>F]AV-1451 meta temporal SUVR | 1.15 (0.08) | 1.21 (0.07) | 1.13 (0.07) | **<0.001** | 1.23(0.04) | 1.11 (0.06) | **<0.001** | 1.13 (0.08) | 1.16 (0.08) | 0.3 |
| Cortical thickness composite measure | 2.75 (0.16) | 2.77 (0.15) | 2.75 (0.16) | 0.6 | 2.78 (0.14) | 2.74 (0.16) | 0.2 | 2.58 (0.15) | 2.83 (0.08) | **<0.001** |
| **Plasma biomarker measures, pg/mL, Mean (SD)** |  |  |  |  |  |  |  |  |  |  |
| Aβ42 | 5.13 (1.62) | 4.95 (1.21) | 5.18 (1.74) | 0.5 | 5.08 (1.39) | 5.15 (1.74) | 0.7 | 5.41(1.76) | 5.00 (1.56) | 0.4 |
| Aβ40 | 81 (23) | 89 (19) | 79 (24) | **0.017** | 84 (29) | 80 (20) | 0.5 | 84 (24) | 80 (23) | 0.8 |
| Aβ42/Aβ40 | 0.063 (0.014) | 0.056 (0.009) | 0.065 (0.015) | **<0.001** | 0.060 (0.011) | 0.064 (0.015) | **0.030** | 0.065 (0.016) | 0.062 (0.013) | 0.2 |
| GFAP | 99 (49) | 124 (37) | 91 (50) | **<0.001** | 102 (54) | 97 (46) | 0.7 | 108 (46) | 95 (50) | 0.10 |
| NfL | 19 (9) | 22 (11) | 18 (8) | 0.11 | 20 (11) | 18 (9) | 0.8 | 24 (12) | 16 (7) | **0.002** |
| p-tau181 | 2.41 (1.47) | 2.81 (1.06) | 2.28 (1.56) | **0.007** | 2.85 (1.69) | 2.17(1.29) | **0.021** | 2.58 (1.25) | 2.35 (1.54) | 0.14 |
| p-tau217 | 0.45 (0.26) | 0.74 (0.29) | 0.35 (0.17) | **<0.001** | 0.57 (0.33) | 0.38 (0.20) | **0.002** | 0.45 (0.22) | 0.45 (0.28) | 0.5 |
| p-tau231 | 9.5 (11.3) | 10.3 (5.2) | 9.3 (12.6) | **0.034** | 12.5 (17.8) | 8.0 (5.0) | **0.037** | 9.2 (4.0) | 9.7 (13.2) | 0.10 |

<sup>a</sup>p-values were calculated using Wilcoxon rank-sum test and fisher's exact test for continuous and categorical variables, respectively. Bold p-values indicate statistical significance.
<sup>b</sup>Area Deprivation Index (ADI) national rank is dichotomized as: Low ADI= 0%–80%; High ADI (i.e. the most disadvantaged) = 80%–100%
Abbreviations: ATN: amyloid-beta, tau, neurodegeneration; PET: positron emission tomography; Aβ42/Aβ40: amyloid-beta42/amyloid-beta40; GFAP: glial fibrillary acidic protein; NfL: neurofilament light chain; p-tau181: phosphorylated-tau181; p-tau217: phosphorylated-tau217; p-tau231:
phosphorylated-tau231; *APOE4*: Apolipoprotein E4; CDR: Clinical Dementia Rating; MMSE: mini-mental status examination score; SUVR: standard uptake value ratio; PiB: Pittsburgh Compound B; SD: standard deviation; A+ (Aβ -PET+): [ $^{11}\text{C}$ ]PiB global SUVR >1.346; A- (Aβ -PET -): [ $^{11}\text{C}$ ]PiB global SUVR ≤ 1.346; T+ (tau-PET+): [ $^{18}\text{F}$ ]AV-1451 meta temporal SUVR >1.18; T- (tau-PET -): [ $^{18}\text{F}$ ]AV-1451 meta temporal SUVR ≤ 1.18; N+ (Neurodegeneration +): cortical thickness composite measure <2.7; N- (Neurodegeneration -): cortical thickness composite measure ≥ 2.7

Between A+ and A- groups, significant differences were found for *APOE4* carrier status (p<0.001), [^11^C]PiB global SUVR measure (p<0.001) and [^18^F]AV-1451 meta temporal SUVR measure (p-value<0.001). The A+ group had a higher percentage of *APOE4* carriers than the A- group (43% in A+ vs 7.1% in the A-). Similarly, between T+ and T- groups a significant difference was found in terms of [^11^C]PiB global SUVR measure (p<0.001) and [^18^F]AV-1451 meta temporal SUVR measure (p-value<0.001). Whereas, between T+ and T- groups the expected difference was seen in terms of *APOE4* carrier status, however the difference was not statistically significant (23% in T+ vs 12% in the T-, p=0.13). No significant difference was found across the “A” and “T” groups in terms of age, sex, race/ethnicity, education level, ADI, mini-mental status examination (MMSE) score, BMI and cortical thickness composite measure. However, a significant difference was seen in terms of age (mean (SD): 80 (7) vs. 75 (5), p=0.002) and cortical thickness composite measure (mean (SD): 2.58 (0.15) vs 2.83 (0.08), p<0.001) between the N+ and N- groups. See **Table 1** for detailed characteristics of the study participants and comparisons across the different neuroimaging-based ATN status groups.

### 3.2. Plasma biomarker concentrations across neuroimaging-based ATN groups

As shown in **Figure 1**, group comparison of log transformed plasma biomarker concentration between the A+ and A- groups showed a statistically significant higher levels of GFAP (p<0.001), p-tau181 (p=0.007), p-tau217 (p<0.001) and p-tau231 (p=0.034) and lower levels of Aβ42/Aβ40 (p<0.001) in the A+ group. Statistically significant higher levels of p-tau181 (p=0.021), p-tau217 (p=0.002) and p-tau231 (p=0.037), and lower level of Aβ42/Aβ40 (p=0.03) was seen in the T+ group compared to the T- group. Only NfL was shown to be significantly higher in the N+ compared with the N- group (p=0.002).

**Figure 1:**
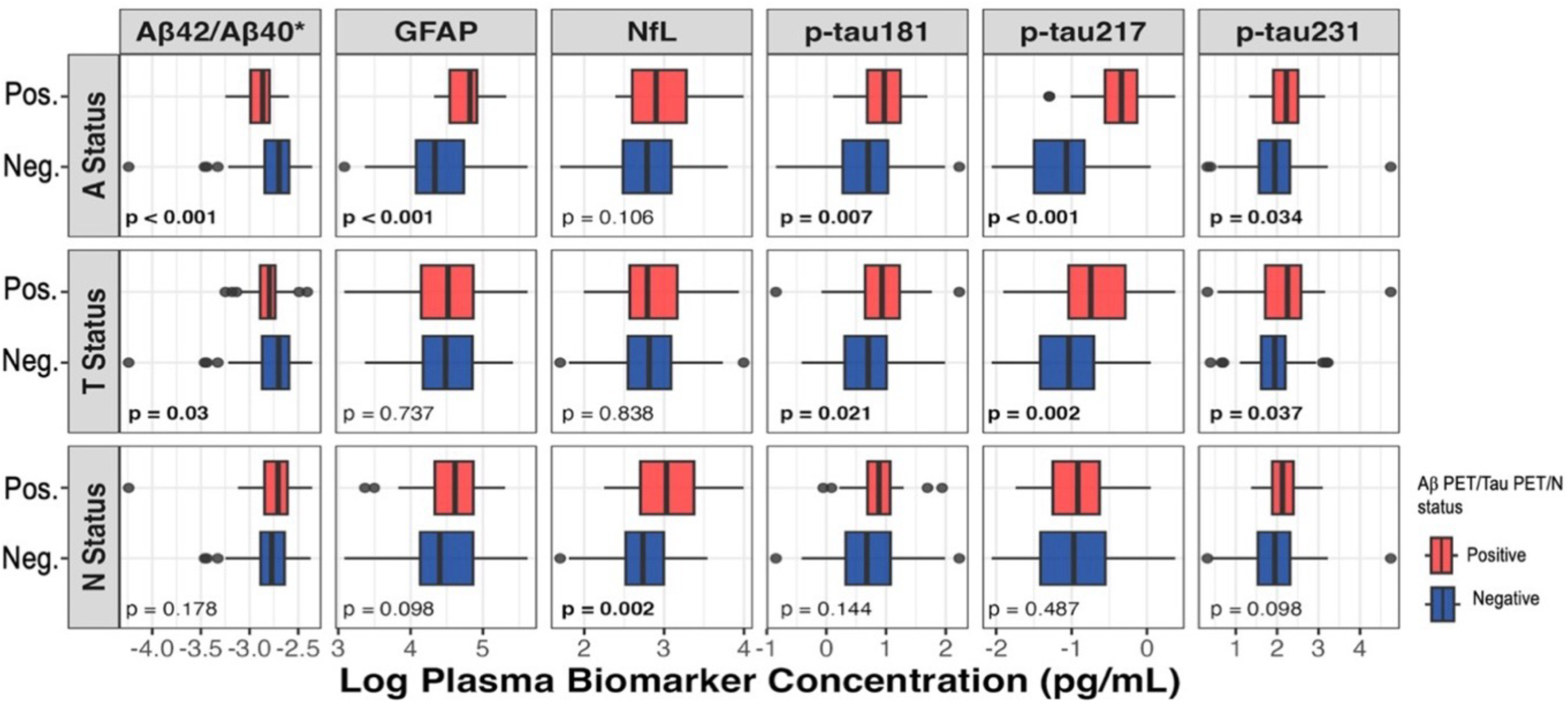
Boxplot comparisons of unadjusted log transformed plasma biomarker concentrations across neuroimaging-based ATN groups. Box ends represent the 25^th^ and 75^th^ percentiles and the horizontal line within each box indicates the median. Whiskers extend to the upper and lower adjacent values. P-values from two-sided student’s t-test are shown. Abbreviations: ATN: amyloid-beta, tau, neurodegeneration: Aβ42/Aβ40- amyloid- beta42/amyloid-beta40; GFAP- glial fibrillary acidic protein; NfL- neurofilament light chain; p- tau181- phosphorylated-tau181; p-tau217- phosphorylated-tau217; p-tau231- phosphorylated- tau231; pg/mL: picogram per milli liter; SUVR: standard uptake value ratio. A+: [^11^C]PiB global SUVR >1.346; T+: [^18^F]AV-1451 meta temporal SUVR >1.18; N+: cortical thickness composite measure <2.7 * Aβ42/Aβ40 measured in units of logarithm of concentration ratio

### 3.3. Association of plasma biomarkers with neuroimaging-based ATN biomarkers

Of the six plasma biomarkers evaluated, Aβ42/Aβ40 and p-tau217 showed significant associations with Aβ-PET status (positive vs negative) in both the unadjusted and adjusted models for AD commonly known risk factors: age, sex, education level, *APOE4* carrier status, and cognitive measure, CDR **(Table 2).** The associations were stronger in the adjusted models: Aβ42/Aβ40 (adjusted odds ratio (AOR)= 1.74*10^-24^, 95% confidence interval (CI) [2.71*10^-46^– 1.12*10^-6^]); p-tau217 (AOR= 3.43*10^3^, 95% CI [126.79–233.13*10^3^]). For plasma p-tau181, the association with Aβ-PET status were statistically significant when adjusted (AOR=1.47, 95% CI [1.02–2.21]). However, no significant association was found between Aβ-PET status and plasma GFAP, NfL, and p-tau231 in either the unadjusted or adjusted models.

**Table 2:** Association between plasma and neuroimaging-based ATN biomarkers.

|  | Unadjusted OR (95% CI) | Adjusted OR (95% CI)<sup>a</sup> |
| --- | --- | --- |
| **A status ~ plasma biomarkers\*** |  |  |
| Aβ42/Aβ40 | **5.14\*10<sup>-23</sup> (4.01\*10<sup>-39</sup> – 1.33\*10<sup>-8</sup>)** | **1.74\*10<sup>-24</sup> (2.71\*10<sup>-46</sup> – 1.12\*10<sup>-6</sup>)** |
| GFAP | 1.01 (1.00–1.02) | 1.01 (1.00–1.02) |
| NfL | 1.04 (1.00–1.09) | 1.05 (0.99–1.12) |
| P-tau181 | 1.25 (0.94–1.69) | **1.47 (1.02–2.21)** |
| p-tau217 | **1.64\*10<sup>3</sup> (117–4.52\*10<sup>4</sup>)** | **3.43\*10<sup>3</sup> (126.79–233.13\*10<sup>3</sup>)** |
| p-tau231 | 1.01 (0.96–1.05) | 1.01 (0.96–1.05) |
| **T status ~ plasma biomarkers\*** |  |  |
| Aβ42/Aβ40 | 6.56*10<sup>-10</sup> (4.75*10<sup>-23</sup>–2.42*10<sup>3</sup>) | 2.79*10<sup>-10</sup> (5.92*10<sup>-22</sup>–5.76*10<sup>5</sup>) |
| GFAP | 1.00 (0.99–1.01) | 1.00 (0.99–1.01) |
| NfL | 1.02 (0.98–1.06) | 1.02 (0.97–1.07) |
| P-tau181 | **1.37 (1.04–1.89)** | **1.50 (1.01– 2.15)** |
| p-tau217 | **19.16 (3.80–121.67)** | **22.24 (3.64–177.85)** |
| p-tau231 | 1.07 (1.00–1.15) | 1.07 (1.00–1.16) |
| **N status ~ plasma biomarkers\*** |  |  |
| Aβ42/Aβ40 | 2.29*10<sup>7</sup> (2.24*10<sup>-6</sup>–1.66*10<sup>21</sup>) | 1.00*10<sup>8</sup> (3.18*10<sup>-11</sup>–4.62*10<sup>23</sup>) |
| GFAP | 1.01 (1.00–1.01) | 1.00 (0.99–1.01) |
| NfL | **1.10 (1.05–1.16)** | **1.09 (1.03–1.15)** |
| P-tau181 | 1.11 (0.82–1.47) | 1.10 (0.76–1.54) |
| p-tau217 | 1.13 (0.23–5.07) | 0.59 (0.083–3.52) |
| p-tau231 | 1.00 (0.94–1.03) | 0.99 (0.93–1.03) |

\* plasma biomarkers measured in picogram per milliliter (pg/mL); <sup>a</sup>Adjusted for age, sex, education level, APOE4 carrier status, Clinical Dementia Rating (CDR)
Abbreviations: ATN: amyloid-beta, tau, neurodegeneration; Aβ42/Aβ40: amyloid-beta42/amyloid-beta40; GFAP: glial fibrillary acidic protein; NfL: neurofilament light chain; p-tau181: phosphorylated-tau181; p-tau217: phosphorylated-tau217; p-tau231: phosphorylated-tau231; OR: odds ratio; CI: confidence interval. A status: Aβ-PET status (positive/negative); T status: tau-PET status (positive/negative); N status: Neurodegeneration status (positive/negative); A status positive: [<sup>11</sup>C]PiB global SUVR >1.346; T status positive: [<sup>18</sup>F]AV-1451 meta temporal SUVR >1.18; N status positive: cortical thickness composite measure <2.7.

With regards to tau-PET status, plasma p-tau181 and p-tau217 showed significant association while adjusted. Similarly, plasma p-tau217 showed the strongest association with tau-PET status (AOR= 22.24, 95% CI 3.64–177.85) followed by plasma p-tau181 (AOR=1.50, 95% CI 1.01–2.15).

Whereas only plasma NfL showed a statistically significant association with neurodegeneration based on cortical thickness measure in both unadjusted and adjusted models with common AD risk factors and cognitive measure (AOR= 1.09, 95% CI 1.03–1.15). No significant association was seen between plasma Aβ42/Aβ40, GFAP, p-tau181, p-tau217, p-tau231 and neurodegeneration based on cortical thickness measure **(Table 2).**

### 3.4. Accuracy of plasma biomarkers in classifying neuroimaging-based ATN status

Summary results of the ROC curves demonstrating the classification accuracies of the plasma biomarkers are shown in **Figure 2**. Plasma p-tau217 showed the highest AUC in distinguishing A+ from A- (AUC= 0.94, 95% CI (0.90–0.98)), followed by plasma Aβ42/Aβ40 (AUC= 0.88, 95% CI (0.78–0.94)) and p-tau181 (AUC= 0.85, 95% CI (0.76–0.93)), while adjusting for age, sex, education level, *APOE4* carrier status and CDR **(Figure 2A).**

**Figure 2:**
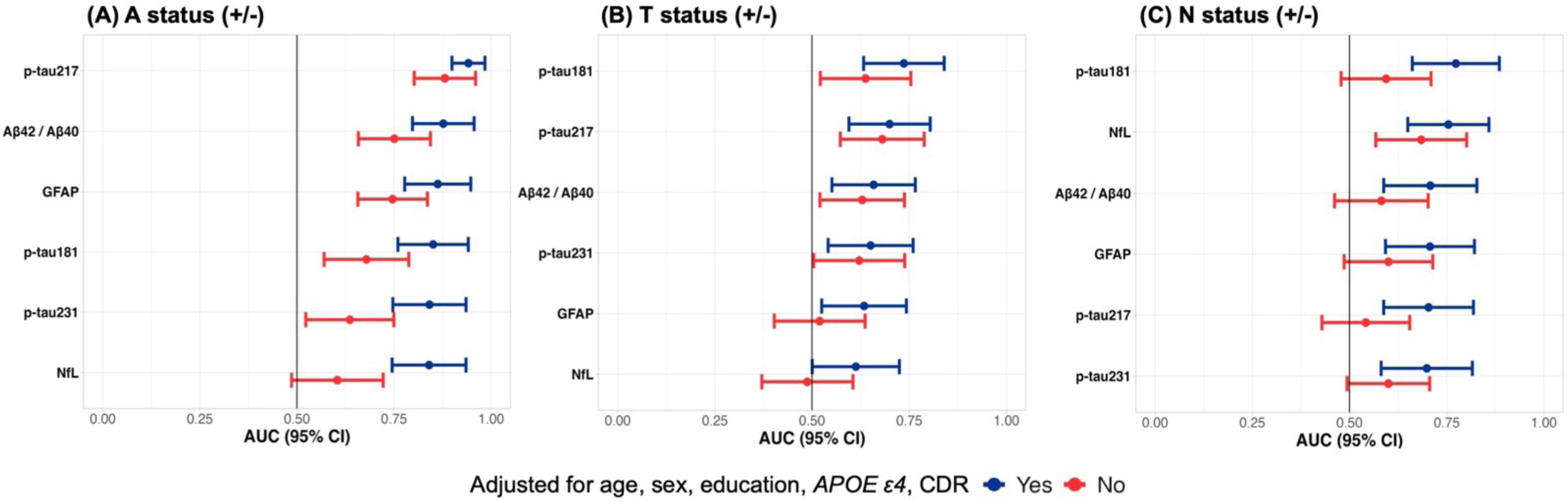
Classification accuracies of plasma biomarkers to identify neuroimaging-based ATN status. Forest plot of area under the receiver operating characteristics curve (ROC AUC) values summary with their corresponding 95% confidence intervals (represented by the error bars and arranged in descending order of the ROC AUC values) illustrating the predictive accuracies of plasma Aβ42/Aβ40, GFAP, NfL, p-tau181, p-tau217, p-tau231 for: **(A)** Aβ-PET, **(B)** tau-PET, **(C)** neurodegeneration, statuses. Abbreviations: Aβ42/Aβ40: amyloid-beta42/amyloid-beta40; GFAP: glial fibrillary acidic protein; NfL: neurofilament light chain; p-tau181: phosphorylated-tau181, p-tau217: phosphorylated- tau217; p-tau231: phosphorylated-tau231; AUC: area under the curve; CI: confidence interval. A status (+/-): Aβ -PET status (positive/negative); T status (+/-): tau-PET status (positive/negative); N status (+/-): Neurodegeneration status (positive/negative); A status positive: [^11^C]PiB global SUVR >1.346; T status positive: [^18^F]AV-1451 meta temporal SUVR >1.18; N status positive: cortical thickness composite measure <2.7.

Similarly, plasma p-tau181 and p-tau217 demonstrated the highest AUC in differentiating T+ from T- individuals (AUC= 0.74, 95% CI (0.64–0.84) and AUC=0.70, 95% CI (0.59–0.80)) respectively, while adjusted **(Figure 2B)**. Whereas NfL and plasma p-tau181 showed the best performance in distinguishing N+ from N- status (AUC=0.75, 95% CI (0.65–0.86) and (AUC= 0.77, 95% CI (0.65–0.87)) respectively, while adjusted **(Figure 2C)**.

Lastly, the results of the sensitivity analysis demonstrated that both the association of the plasma biomarkers with neuroimaging-based ATN biomarkers and their classification accuracy of neuroimaging-based ATN status were robust to the adjustment for ADI **(Supplementary Table 2)** and BMI **(Supplementary Tables 3).** Only NfL and plasma p-tau181 classification accuracies in distinguishing N+ from N- status slightly improved when BMI was included in the adjusted model (AUC=0.79 and AUC=0.80, respectively) **(Supplementary Table 3).**

### 3.5. Voxel-wise associations of A**β**-PET SUVR with plasma biomarkers

We performed voxel-wise linear regression analysis between plasma biomarkers and Aβ-PET SUVR images. We found a significant positive association between Aβ-PET and plasma p- tau217 (**Figure 3A**) in AD-related regions, such as fusiform, inferior temporal and precuneus; in regions such as inferior and middle temporal gyrus with GFAP (**Supplementary** Figure 1), and a small cluster in the inferior temporal region with NfL (**Figure 3A**). Plasma Aβ42/Aβ40 was negatively associated with Aβ-PET in Aβ deposition regions, such as frontal lobe and anterior cingulate (**Figure 3A**).

**Figure 3:**
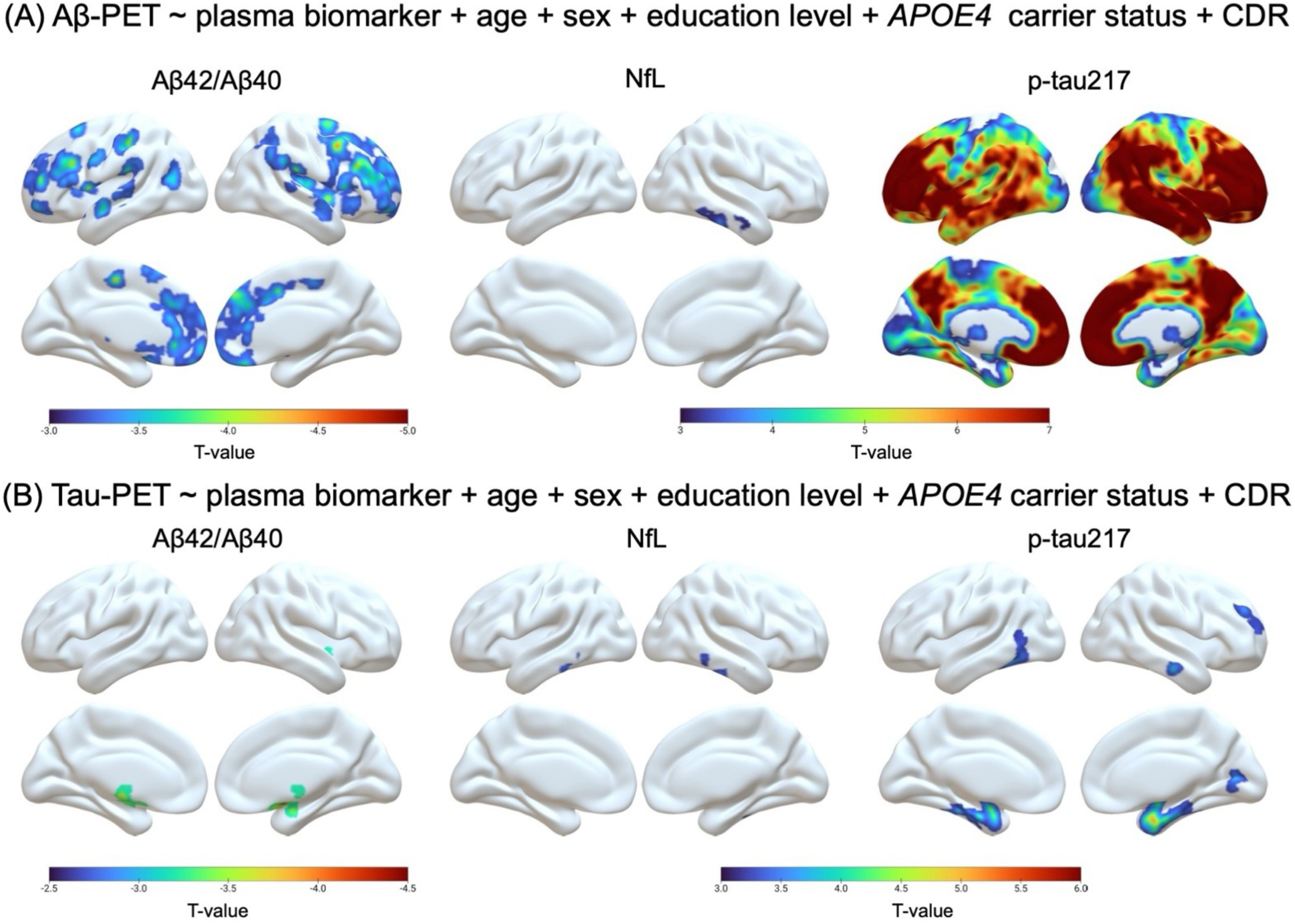
Voxel-wise associations between plasma biomarkers and. **A**β-PET (Panel A) and tau-PET (Panel B); after adjusting for age, sex, education level, *APOE4* carrier status, and CDR. All results were adjusted for multiple comparisons using random field theory with a voxel threshold of p<0.001. Abbreviations: PET-positron emission tomography; *APOE4*: Apolipoprotein E4; CDR- clinical dementia rating; SUVR-standard uptake value ratio; Aβ42/Aβ40-amyloid-beta42/amyloid-beta 40; NfL- neurofilament light chain; p-tau217- phosphorylated-tau217.

### 3.6. Voxel-wise associations of tau-PET SUVR with plasma biomarkers

We performed voxel-wise linear regression analysis between plasma biomarkers and tau-PET SUVR images. We found a significant positive association between tau-PET and plasma p- tau217 (**Figure 3B**) in early Braak regions (Braak I and II); and in small clusters with plasma Aβ42/Aβ40, NfL (**Figure 3B**), p-tau181 and p-tau231 **(Supplementary** Figure 1).

## 4. Discussion

In this population-based study composed of mostly cognitively normal older adults from a socioeconomically disadvantaged area, we investigated six plasma biomarkers in parallel (i.e., Aβ42/Aβ40, GFAP, NfL, p-tau181, p-tau217, and p-tau231), regarding predicting Aβ (A) and tau (T) pathologies and neurodegeneration (N) on neuroimaging. Thus, we evaluated their potential utility as ATN markers of AD.

The major findings were: (i) Of the six plasma biomarkers, both plasma Aβ42/Aβ40 ratio and p- tau217 showed strong associations with Aβ-PET status independent of common risk factors and a cognitive measure: age, sex, education level, *APOE4* carrier status and CDR. Plasma p- tau181 showed a significant association with Aβ-PET status only when combined with the aforementioned AD common risk factors and cognitive measure. (ii) Plasma p-tau181 and p- tau217 were consistently associated with tau-PET status i.e. independent of common risk factors and cognitive measure. (iii) Similarly, on a voxel scale, p-tau217 was strongly associated with neocortical Aβ deposition. (iv) Only plasma NfL was associated with neurodegeneration status as determined by MRI-based cortical thickness composite measure. (v) Notably, plasma p-tau217 demonstrated the highest classification performance for distinguishing individuals with Aβ-PET positive from negative profiles, followed by Aβ42/Aβ40. (vi) Whereas plasma p-tau181 and p-tau217 for tau status, and comparably p-tau181 and NfL for neurodegeneration status showed the best performance. The classification performances of all plasma biomarkers were improved when combined with age, sex, education level, *APOE4* carrier status and CDR.

Recent advancements in plasma biomarker assays have enabled more accessible and non-invasive prediction of brain Aβ deposition. Plasma Aβ42/Aβ40 is one of the biological markers to change during the early preclinical stage of AD [8,9,46]. Unlike the individual levels of Aβ42, the Aβ42/Aβ40 ratio has consistently shown a strong concordance with brain amyloidosis, likely because the ratio may normalize differences related to biological variations, circadian rhythms or sample processing [47,48]. In line with our observation, strong inverse correlation has been widely reported previously among individuals without dementia [8,9,49–53]. In studies done by Schindler et.al. and Li et.al., similar high classification accuracy results (ROC AUC ranging 0.88- 0.94) were found [8,51], despite immunoprecipitation-mass spectrometry methods having been used in those studies, compared to immunoassays used in the current study. These findings highlight the potential of plasma Aβ42/Aβ40 as a reliable surrogate marker of brain amyloidosis enabling efficient population-wide screening.

Among the three p-tau epitopes assessed in our study, plasma p-tau217 was found to be the strongest predictor of brain Aβ deposition with the best classification performance (ROC AUC=0.94), even higher than plasma Aβ42/Aβ40. Similar results have been reported in several studies where plasma p-tau217 has been correlated with brain amyloidosis in early disease stages [14,15,17,18,23,27]. For instance, a study done in the ALFA+ cohort of individuals with pre-clinical AD, showed that plasma p-tau217 had the strongest association with Aβ-PET in early accumulating regions with 89% classification accuracy [14]. Moreover, a study done by Schindler et.al. showed that plasma p-tau217 measures, either individually or in combination, had the strongest association with all AD outcome measures [26]. In our study, compared to plasma p-tau217, p-tau181 showed a weaker association and lower classification accuracy for Aβ-PET status. This is in line with a head-to-head comparative study done by Mendes et.al. [54], and Mielke et.al. [55] revealing that plasma p-tau217 might be more efficient than plasma p-tau181 in identifying Aβ-PET positivity.

Previous studies have extensively explored biologically plausible molecular and cellular mechanisms to understand the underlying relationship between the various isoforms of phosphorylated tau and brain Aβ pathology [56–60]. Evidence indicates that Aβ plays an upstream role in AD progression by promoting tau phosphorylation [56]. Aβ plaques are linked to an increase in the release of axonal hyperphosphorylated tau, which subsequently leads to the formation and progression of tau neurofibrillary tangle pathology [59]. This apparent Aβ- triggered release of soluble phosphorylated tau forms in blood starts at one of the earliest stages of the AD continuum. Overall, our finding highlights the robustness of plasma p-tau217 to identify early brain amyloidosis in dementia-free community-dwelling individuals and can serve as a stand-alone biomarker for screening of high-risk individuals for early intervention as well as for participation in AD clinical trials and disease-modifying therapies, while reducing the need for the use of invasive (e.g., CSF) or neuroimaging biomarkers.

Tau deposition is another important hallmark of AD neuropathology. Our findings of significant association between both plasma p-tau181 and p-tau217 and tau-PET status are consistent with results from other studies [19,20,23–25]. Interestingly, both biomarkers, however, showed weaker association and lower classification accuracy for tau-PET than for Aβ-PET. This adds to the understanding that these plasma p-tau isoforms, closer to the N-terminus of the tau protein, become more closely related to tau tangles, identified by PET, in later stages of AD [23,61]. For instance, a study by Ferreira et.al. demonstrated stronger correlation between plasma p-tau217 and tau-PET signals in cognitively impaired individuals compared to those without impairment [23]. A possible explanation for this could be that tau-PET imaging may not reliably detect smaller amounts of tau aggregates in the brain making it less sensitive to the earliest and most subtle tau pathology [62]. Another factor to consider is that the soluble biochemical pools of tau, with epitopes closer to the N-terminus of the tau protein, are affected – and are detectable – earlier than the microtubule binding domain (MTB), the aggregating portion of the tau molecule, closer to the C-terminus [63]. Despite these limitations, we propose that plasma p-tau181 and p- tau217 could serve as potential screening tools for brain tau pathology.

Furthermore, for “N” status, NfL was the only biomarker we found to demonstrate significant association with a classification accuracy of 75% for detecting neurodegeneration based on cortical thickness measures in cognitively normal individuals. This supports the growing evidence that plasma NfL reflects neuronal damage and could serve as a proxy for imaging-based neurodegeneration markers in various neurodegenerative conditions, including AD [28,64]. A prior study done by Matsson et al. reported strong associations between plasma NfL and hippocampal atrophy, as well as cortical thickness–both indicators of neurodegeneration in AD [65]. Additionally, studies conducted on individuals with AD-related genetic mutations reported that blood NfL can predict the onset of AD more than a decade before clinical manifestations [65,66]. Although plasma NfL is not AD-specific, given our findings, we propose its potential usefulness as a non-invasive and more accessible screening biomarker for neurodegeneration. Subsequent studies will evaluate AD-type neurodegeneration markers such as brain-derived tau [67–69].

Finally, it is worth noting that in our study we found no statistically significant association between plasma GFAP and the “T” and “N” neuroimaging biomarkers, and between p-tau231 and the “A” and “N” neuroimaging biomarkers. This partly aligns with a study by Pereira et.al. on the Swedish BioFINDER-2 cohort, that reported no correlation between plasma GFAP and tau aggregation as measured by PET in cognitively normal individuals [11]. Additionally, plasma GFAP was found to be only weakly associated with hippocampal volume [70]. In contrast, other studies have reported significant associations between both plasma GFAP and p-tau231; specially with Aβ-PET measures in early stages of AD [10,14,21,23]. These inconsistencies highlight the need for further research to clarify the roles of these plasma biomarkers along the AD continuum.

Our study has several strengths. First, the MYHAT-NI cohort recruited participants directly from an active, randomly recruited population-based cohort which reduces the selection bias often present in clinic-based or convenience sampling. This improves the applicability of our findings to a wider population of older adults. However, because the cohort consisted largely of individuals identifying as non-Hispanic white, further validation is needed across diverse racial and ethnic groups. Second, the study was conducted in socioeconomically disadvantaged small-town communities. This ensures our study can shed light on the limited information we have on how AD plasma biomarkers perform in populations living in under-resourced, small-town areas. Third, we studied the performance of several novel AD plasma biomarkers in parallel obtained from the same individuals using well-established neuroimaging reference standards. Fourth, we accounted for several known AD risk factors and cognitive measure, enhancing robustness of our findings. A key limitation of the study is the lack of additional SES and medical variables. Consideration of other structural and social determinants of health and health-related factors, such as chronic kidney disease and creatinine level (that are found to be related to certain plasma biomarkers levels [71]), could provide a more comprehensive understanding of ATN biomarkers associations and performance in diverse populations and health conditions and should be incorporated in future work. Furthermore, the small number of CDR = 0.5 participants in the present study (11 out of 113) limits insights gained into impact of early cognitive decline to the outcomes.

In conclusion, this study has identified promising plasma biomarkers to identify AD ATN pathologies in dementia-free individuals. Plasma p-tau217 emerged as a robust stand-alone predictor of both amyloid and tau statuses, with plasma Aβ42/Aβ40, p-tau181, and NfL also demonstrating strong predictive capabilities for “A”, “T” and “N” statuses, respectively. Our findings underscore the potential of using a combination of plasma biomarkers and known risk factors and cognitive measure to enhance classification accuracy and make them valuable tools for selecting individuals for clinical trials and disease-modifying therapies. Importantly, our results demonstrate that, in this community-based cohort, cortical thickness was not related to amyloid pathology perhaps due to the latter becoming abnormal earlier. Moreover, the findings including ATN profile distributions agree with multiple published papers showing strong association and classification accuracies of leading plasma biomarkers such as p-tau217 for AD. Together, these results concord with other investigations performed in cohorts recruited from different settings including in clinics and populations of other demographics, supporting the use of plasma biomarkers for AD detection and monitoring in community-based settings.

## Data Availability

All data produced in the present study are available upon reasonable request to the authors. Data requests will be promptly reviewed and processed in accordance with University of Pittsburgh procedures and the IRB stipulations.

## Acknowledgments

We are grateful for our study participants, their families, caregivers, staff of the MYHAT-NI and MYHAT studies.

## Author contribution

MTD, XZ, BES, CES, and TKK contributed to the study’s conception and design. BES and MG led the MYHAT-NI cohort study establishment including participant recruitment, cognitive evaluation, neuroimaging measures, among others. AP, JLD, PCLF and TAP led statistical analyses and produced the figures. PCLF, BB, GP, VLV, ADC and TAP contributed to neuroimaging data collection and analysis. MIK led *APOE* genotyping. MTD, AP, PCLF, BES, CES, XZ and TKK were major contributors in writing the initial manuscript draft, which was subsequently critically reviewed by all authors. All authors contributed to and approved the final version of the manuscript.

## Ethics declarations

### Ethics approval and consent to participate

The MYHAT-NI study was performed under written informed consent and approved by the University of Pittsburgh Institutional Review Board (STUDY19020264). This study was performed in accordance with the Declaration of Helsinki.

## Consent for publication

All participants gave written informed consent, and all study procedures were approved by the Institutional Review Board of the University of Pittsburgh.

## Abbreviations

Aβ: Amyloid-beta
AD: Alzheimer’s disease
*APOE4*: Apolipoprotein E4
ATN: Amyloid-beta, tau, neurodegeneration
AUC: Area under the curve
CDR: Clinical dementia rating
CSF: Cerebrospinal fluid
CV: Coefficient of variation
GFAP: Glial fibrillary acidic protein
MCI: Mild cognitive impairment
MRI: Magnetic resonance imaging
MYHAT: Monongahela Youghiogheny Healthy Aging Team
MYHAT NI: Monongahela Youghiogheny Healthy Aging Team-Neuroimaging
NfL: Neurofilament light chain
NIA-AA: National Institute on Aging-Alzheimer’s Association
PET: Positron emission tomography
PiB: Pittsburgh compound-B
p-tau: Phosphorylated-tau
Simoa: Single-molecule array
SUVR: Standardized uptake value ratio

