## Supplementary Table 1 for "Plasma biomarkers identify brain ATN abnormalities in a dementia-free population-based cohort"

**Supplementary Table 1: Neuroimaging-based ATN profile distribution of the MYHAT-NI cohort**

| **ATN profiles** | **N (%)** |
| --- | --- |
| A-T-N- | 45 (40%) |
| A- T- N+ | 20 (18%) |
| A- T- N+ | 14 (12%) |
| A- T+ N+ | 6 (5%) |
| A+ T-N- | 6 (5%) |
| A+ T- N+ | 3 (3%) |
| A+ T+ N- | 15 (13%) |
| A+ T+ N+ | 4 (4%) |

Abbreviations: ATN: amyloid-beta, tau, neurodegeneration; A+ (Aβ-PET+): [^11^C]PiB global SUVR >1.346; A- (Aβ-PET -): [^11^C]PiB global SUVR <=1.346; T+ (tau-PET+): [^18^F]AV-1451 meta temporal SUVR >1.18; T- (tau-PET -): [^18^F]AV-1451 meta temporal SUVR <=1.18; N+ (Neurodegeneration +): cortical thickness composite measure <2.7; N- (Neurodegeneration -): cortical thickness composite measure >= 2.7
