## Supplementary Table 2 for "Plasma biomarkers identify brain ATN abnormalities in a dementia-free population-based cohort"

**Supplementary Table 2. Association of plasma and neuroimaging biomarkers and their classification accuracy**

|  | **Unadjusted OR (95% CI)** | **Unadjusted AUC** | **Adjusted OR (95% CI) ^a^** | **Adjusted AUC^a^** |
| --- | --- | --- | --- | --- |
| **A status ~ plasma biomarkers*** |  |  |  |  |
| Aβ42/Aβ40 | **5.14*10^-23^ (4.01*10^-39^ – 1.33*10^-8^)** | 0.75 | **1.81*10^-24^ (1.56*10^-42^ – 0.25*10^-3^)** | **0.88** |
| GFAP | 1.01 (1.00–1.02) | 0.75 | 1.01 (1.00–1.02) | 0.87 |
| NfL | 1.04 (1.00–1.09) | 0.60 | 1.05 (0.98–1.11) | 0.84 |
| P-tau181 | 1.25 (0.94–1.69) | 0.68 | **1.55 (1.05–2.44)** | **0.86** |
| p-tau217 | **1.64*10^3^ (117–4.52*10^4^)** | 0.88 | **1.18*10^4^ (252.12–199.50*10^4^)** | **0.95** |
| p-tau231 | 1.01 (0.96–1.05) | 0.64 | 1.01 (0.96–1.05) | 0.84 |
| **T status ~ plasma biomarkers*** |  |  |  |  |
| Aβ42/Aβ40 | 6.56*10^-10^ (4.75*10^-23^–2.42*10^3^) | 0.63 | 6.13*10^-7^ (9.54*10^-21^–1.93*10^7^) | 0.66 |
| GFAP | 1.00 (0.99–1.01) | 0.52 | 1.00 (0.99–1.01) | 0.64 |
| NfL | 1.02 (0.98–1.06) | 0.49 | 1.02 (0.97–1.07) | 0.64 |
| P-tau181 | **1.37 (1.04–1.89)** | **0.64** | **1.56 (1.12– 2.29)** | **0.74** |
| p-tau217 | **19.16 (3.80–121.67)** | **0.68** | **20.99 (3.43–170.11)** | **0.70** |
| p-tau231 | 1.07 (1.00–1.15) | 0.62 | 1.07 (1.00–1.17) | 0.66 |
| **N status ~ plasma biomarkers*** |  |  |  |  |
| Aβ42/Aβ40 | 2.29*10^7^ (2.24*10^-6^–1.66*10^21^) | 0.58 | 1.74*10^9^ (3.30*10^-6^–1.85*10^25^) | 0.71 |
| GFAP | 1.01 (1.00–1.01) | 0.60 | 1.00 (0.99–1.01) | 0.70 |
| NfL | **1.10 (1.05–1.16)** | **0.68** | **1.09 (1.03–1.16)** | **0.75** |
| P-tau181 | 1.11 (0.82–1.47) | 0.59 | 1.09 (0.75–1.54) | 0.77 |
| p-tau217 | 1.13 (0.23–5.07) | 0.54 | 0.57 (0.076–3.44) | 0.70 |
| p-tau231 | 1.00 (0.94–1.03) | 0.60 | 0.99 (0.93–1.03) | 0.69 |

^*^plasma biomarkers measured in picogram per milliliter (pg/mL); **^a^**Adjusted for age, sex, education level, *APOE4* carrier status, Clinical Dementia Rating (CDR), Area Deprivation Index (ADI)

Area under the receiver operating characteristics curve (ROC AUC) values show the predictive accuracies of plasma Aβ42/Aβ40, GFAP, NfL, p-tau181, p-tau217, p-tau231 for: **(A)** Aβ-PET, **(B)** tau-PET, **(C)** neurodegeneration, statuses.

Abbreviations: ATN: amyloid-beta, tau, neurodegeneration; Aβ42/Aβ40: amyloid-beta42/amyloid-beta40; GFAP: glial fibrillary acidic protein; NfL: neurofilament light chain; p-tau181: phosphorylated-tau181; p-tau217: phosphorylated-tau217; p-tau231: phosphorylated-tau231; OR: odds ratio; CI: confidence interval. A status: Aβ-PET status (positive/negative); T status: tau-PET status (positive/negative); N status: Neurodegeneration status (positive/negative); A status positive: [^11^C]PiB global SUVR >1.346; T status positive: [^18^F]AV-1451 meta temporal SUVR >1.18; N status positive: cortical thickness composite measure <2.7.
