## Supplementary Table 3 for "Plasma biomarkers identify brain ATN abnormalities in a dementia-free population-based cohort"

**Supplementary Table 3. Association of plasma and neuroimaging biomarkers and their classification accuracy**

|  | **Unadjusted OR (95% CI)** | **Unadjusted AUC** | **Adjusted OR (95% CI) ^a^** | **Adjusted AUC^a^** |
| --- | --- | --- | --- | --- |
| **A status ~ plasma biomarkers*** |  |  |  |  |
| Aβ42/Aβ40 | **5.14*10^-23^ (4.01*10^-39^ – 1.33*10^-8^)** | **0.75** | **2.84*10^-20^ (7.13*10^-40^ – 3.7*10^-8^)** | **0.87** |
| GFAP | 1.01 (1.00–1.02) | 0.75 | 1.01 (1.00–1.02) | 0.85 |
| NfL | 1.04 (1.00–1.09) | 0.60 | 1.04 (0.97–1.11) | 0.85 |
| P-tau181 | 1.25 (0.94–1.69) | **0.68** | **1.41 (0.96–2.15)** | **0.83** |
| p-tau217 | **1.64*10^3^ (117–4.52*10^4^)** | **0.88** | **4.07*10^3^ (127.83–362.08*10^3^)** | **0.94** |
| p-tau231 | 1.01 (0.96–1.05) | 0.64 | 1.01 (0.94–1.05) | 0.85 |
| **T status ~ plasma biomarkers*** |  |  |  |  |
| Aβ42/Aβ40 | 6.56*10^-10^ (4.75*10^-23^–2.42*10^3^) | 0.63 | 3.02*10^-8^ (1.75*10^-22^–2.59*10^6^) | 0.66 |
| GFAP | 1.00 (0.99–1.01) | 0.52 | 1.00 (0.99–1.01) | 0.63 |
| NfL | 1.02 (0.98–1.06) | 0.49 | 1.03 (0.98–1.09) | 0.62 |
| P-tau181 | **1.37 (1.04–1.89)** | **0.64** | **1.40 (1.01– 2.03)** | **0.72** |
| p-tau217 | **19.16 (3.80–121.67)** | **0.68** | **17.91 (2.38–167.24)** | **0.67** |
| p-tau231 | 1.07 (1.00–1.15) | 0.62 | 1.06 (1.00–1.15) | 0.66 |
| **N status ~ plasma biomarkers*** |  |  |  |  |
| Aβ42/Aβ40 | 2.29*10^7^ (2.24*10^-6^–1.66*10^21^) | 0.58 | 2.31*10^6^ (1.54*10^-9^–5.53*10^22^) | 0.74 |
| GFAP | 1.01 (1.00–1.01) | 0.60 | 1.00 (0.99–1.02) | 0.73 |
| NfL | **1.10 (1.05–1.16)** | **0.68** | **1.1 (1.04–1.17)** | **0.79** |
| P-tau181 | 1.11 (0.82–1.47) | 0.59 | 1.24 (0.85–1.79) | 0.80 |
| p-tau217 | 1.13 (0.23–5.07) | 0.54 | 1.27 (0.14–11.16) | 0.73 |
| p-tau231 | 1.00 (0.94–1.03) | 0.60 | 0.99 (0.93–1.03) | 0.73 |

^*^plasma biomarkers measured in picogram per milliliter (pg/mL); **^a^**Adjusted for age, sex, education level, *APOE4* carrier status, Clinical Dementia Rating (CDR), Body Mass Index (BMI)

Area under the receiver operating characteristics curve (ROC AUC) values show the predictive accuracies of plasma Aβ42/Aβ40, GFAP, NfL, p-tau181, p-tau217, p-tau231 for: **(A)** Aβ-PET, **(B)** tau-PET, **(C)** neurodegeneration, statuses.

Abbreviations: ATN: amyloid-beta, tau, neurodegeneration; Aβ42/Aβ40: amyloid-beta42/amyloid-beta40; GFAP: glial fibrillary acidic protein; NfL: neurofilament light chain; p-tau181: phosphorylated-tau181; p-tau217: phosphorylated-tau217; p-tau231: phosphorylated-tau231; OR: odds ratio; CI: confidence interval. A status: Aβ-PET status (positive/negative); T status: tau-PET status (positive/negative); N status: Neurodegeneration status (positive/negative); A status positive: [^11^C]PiB global SUVR >1.346; T status positive: [^18^F]AV-1451 meta temporal SUVR >1.18; N status positive: cortical thickness composite measure <2.7.
