## Supplementary Figure 1 for "Plasma biomarkers identify brain ATN abnormalities in a dementia-free population-based cohort"

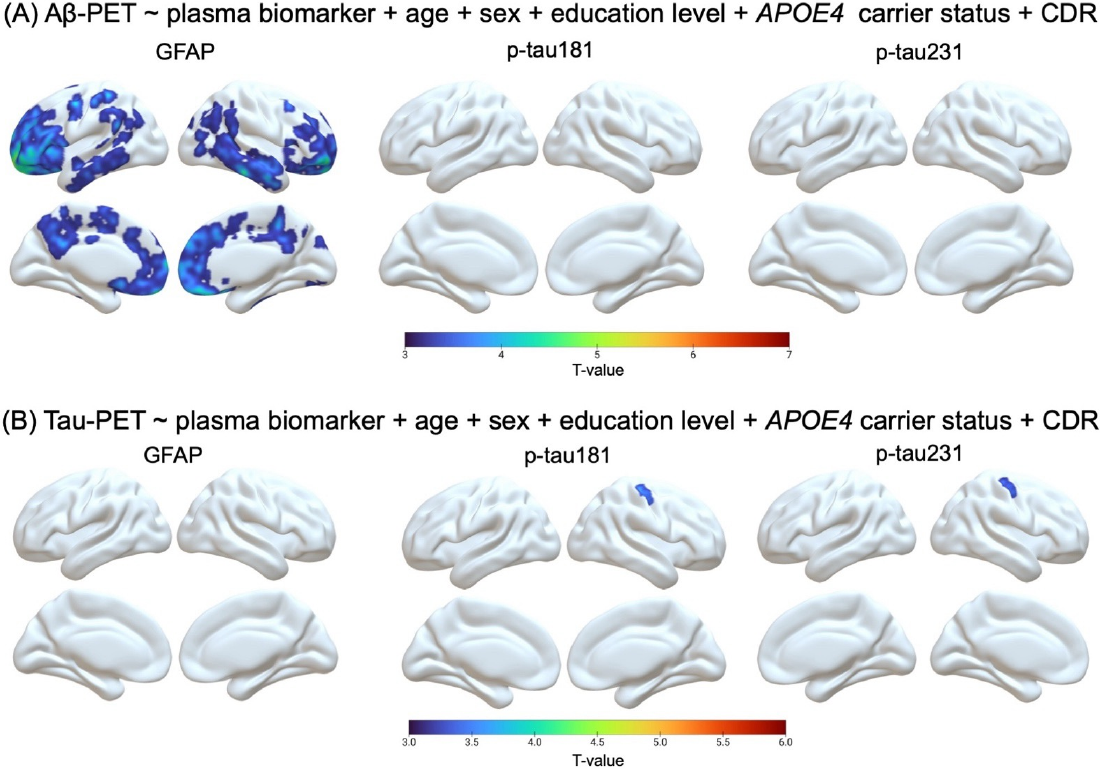

**Supplementary Figure 1:** **Voxel-wise associations between plasma biomarkers and Aβ-PET (Panel A) and tau-PET (Panel B); after adjusting for age, sex, education level, *APOE4* carrier status, and CDR.** All results were adjusted for multiple comparisons using random field theory with a voxel threshold of p<0.001. Abbreviations: Aβ: amyloid-beta; PET: positron emission tomography; *APOE4*: Apolipoprotein E4; CDR: clinical dementia rating; GFAP: glial fibrillary acidic protein; p-tau181: phosphorylated-tau181; p-tau231: phosphorylated-tau231.
